# Uncovering spatial-temporal patterns in mortality counts from pulmonary embolism in US counties between 2005 to 2022

**DOI:** 10.64898/2026.04.16.26351045

**Authors:** Ogutu B. Osoro, Diego F Cuadros

## Abstract

Pulmonary embolism (PE) is a sudden blockage of lung arteries, usually caused by a blood clot that travels from the deep veins of the legs. As the world becomes more sedentary and lifestyle diseases emerge, deaths from PE are expected to rise in the next 20 years. For instance, the United States records annual deaths of 60 per 100,000 people. The degree to which these deaths are affected by demographic, socioeconomic and environmental predisposing factors as well as how they vary across time and space remains an open science question. In this paper, we conduct a detailed statistical and spatial-temporal study PE mortality counts across US counties from 2005 to 2022. Our study shows that study shows that PE mortality is not randomly distributed in space and time but concentrated in most counties in Arkansas, Mississippi, Kansas, Missouri, Oklahoma, Louisiana, Nebraska, Tennessee, and Texas. We also established that age is a statistically significant predictor (mean coefficient of 0.52) of PE mortality especially in counties of Mississippi, Kansas, Missouri, Tennessee, Illinois, Kentucky, Texas and Virginia. Our results thus provide empirical support for prioritizing regionally targeted PE prevention policies. Furthermore, the adopted county-level analysis uncovered granular geographic patterns that are usually obscured in state or national level analysis. Our study thus provides actionable evidence to support geographically tailored strategies aimed at reducing mortality by pinpointing counties with consistently elevated PE mortality risk at different timescales.

## 1. Introduction

Pulmonary Embolism (PE) is a condition where a blood clot blocks an artery in the lung thus stopping the flow of blood. PE is part of broader Venous Thromboembolism (VTE) disease where blood clots starts in the veins (Zöller et al., 2012). VTE may also occur within deep vein in the body such as legs often due to inactivity (e.g. bed rest, long flights) injury or surgery. When this clot breaks off and travel upwards to the heart and passes to pulmonary artery it can block the arteries to the lungs, thus resulting to decrease in oxygen levels, right-heart strain, collapse, cardiac arrest and death in some cases. Studies have indicated that PE is a life-threatening condition and the third leading cause of cardiovascular death after coronary artery diseases and stroke (Konstantinides et al., 2014).

It is estimated that PE is responsible for nearly 5 million deaths annually (Nopp et al., 2025). Incidences of PE varies from country to country, and it causes significant morbidity, mortality and economic burden especially in developed countries (Alvaro-Meca et al., 2018). In the United States for instance, PE causes approximately 60 deaths per 100,000 person each year (Zhang et al., 2024). The PE mortality is projected to increase in the next 20 years driven mainly by the rise of predisposing risk factors (Couturaud et al., 2021). First, the world is becoming more sedentary and overweight (Ng et al., 2025) with countries like United States recording 40.3% obesity prevalence among the adult population (Emmerich et al., 2024). Secondly, the incidences and prevalence of other predisposing risk factors such as cancer, trauma, surgery, hormonal therapy and oral contraceptives are on the rise (Aaron et al., 2019). Lastly, international travel (over 4 billion airline passengers in 2025) is also increasing. These developments are expected to add to the overall risk of PE.

Generally, the growth in world population is expected to increase the incidence of PE with countries such as China already witnessing the impact. From 2007 to 2016 alone, the number of PE related hospitalizations in China increased from 3.2 to 17.5 per 100,000 persons (Aaron et al., 2019). The general trend is similar in the United States with up to 300,000 deaths annually (Mehta et al., 2025). Studying the temporal trends in PE mortality is necessary to assist in driving a more data driven and focused healthcare policy interventions. Importantly, it is critical to understand the mortality trends among different age groups, sex and races.

A detailed analysis of PE mortality counts data has shown variations based on age, sex, and race. Indeed, PE mortality risk increase by age especially beyond 70 years (Cash et al., 2022). Moreover, females are at higher risks of PE due to usage of contraceptives and other birth control methods that alters their hormonal balance (Heikinheimo et al., 2022; Santos et al., 2022; Thachil et al., 2022). Attempts to understand PE mortality variation by race have been done with the results showing that Black patients are nearly two times more likely to die from the condition compared to White patients (Afifi et al., 2024). The degree to which these deaths vary between sex, age and racial group across time and space remains an open science question. Limited studies have mostly focused on one segment and at higher spatial level such as state. However, there is need to conduct a detailed statistical and spatial analysis of PE mortality counts across decadal timescales and granular spatial level such as counties.

In addition, there is need to investigate the relationship in variation of PE mortality rates across the geographical scale as it might be driven by external socioeconomic and environmental factors that are common to specific neighborhoods and the environment. For instance, the built-up environment influences the level of physical activity among people in a particular neighborhood. Areas with poor walkability index are likely to encourage more sedentary lifestyles that translates to physical inactivity thus increasing poor blood circulation and likelihood of clots among residents (Baobeid et al., 2021; Kunutsor and Laukkanen, 2024). Similarly, environmental factors such as concentration of Particulate Matter below 2.5 nm (PM_2.5_) and poor air quality can lead to systematic inflammation when inhaled and enters the lungs (Hantrakool et al., 2022; Pei et al., 2025). The inflammation can result into coagulation, platelet activation and fibrin formation that raises the risk of deep vein thrombosis.

Furthermore, socioeconomic determinants such as poverty rate may influence the likelihood of developing blood clots and risk of death after the clot. High-poverty populations tend to have high incidence of PE predisposing conditions such as obesity and diabetes that increase the overall risk. Indeed, PE is a time-sensitive condition whose risk of death increases the longer the clot remains untreated. However, high-poverty neighborhood populations are likely to have limited primary care visits, limited access to imaging, delayed diagnosis and treatment.

These socioeconomic and environmental factors are temporal and spatial in nature. The level at which each or jointly contribute to PE mortality rates across time and space in the United States require further examination. To this end, we use PE mortality count data from the Centers for Disease Control and Prevention (CDC) from 2005 to 2022, to conduct a detailed statistical and geospatial analysis to answer the following key research questions.

1. How do pulmonary embolism (PE) mortality counts vary over time across sex, age groups, and racial populations in the United States?
2. To what extent do demographic, socioeconomic, and environmental factors explain spatial variation in PE mortality across U.S. counties?
3. What are the spatial and temporal patterns of PE mortality hotspots across U.S. counties, and how does the distributions differ by period and sex?

## 2. Methodology

In this section, we first define our spatial-temporal theoretical model framework used in the study. We then apply the theoretical model to analyze the mortality trends, predictions and emerging hotspots from PE across US counties.

### 2.1. Theoretical framework

Given a time series dataset of a given outcome (e.g. mortality counts) grouped by independent categorical variables such as sex, age group or race for each of geographical units, a chi-square test (*χ*^2^) of independence can be done using equation (1) to evaluate if the outcome distribution by the categories is the same across a geographical space.

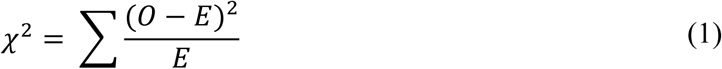

Where *0*, is the observed outcome from the data and *E*, the expected count under independence. Next, considering the outcome as time series, multiple count data at year *t* (i.e. *Y*_*t*_), based on categorical variables (*a*_*t*_) can be observed during the study period as defined by equation (2).

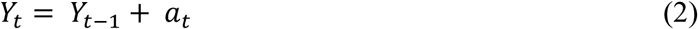

For time series analysis, it is important to establish whether the values of interest are increasing or decreasing over time. Stationarity tests utilizing mean (*μ*) and variance (*σ*) allows for detecting positive or negative changes within a given a time frame. If the statistical moments (*μ* or *σ*) remains unchanged each year, the data is weakly stationary.

Next, an Autocorrelation Function (ACF), *ρ* can be fitted in the time series data to detect cases of stationarity or non-stationarity using equation (3). Calculating autocorrelation of the adjacent observations for each time steps (e.g. every year) lead to series of result, *lag-1* that supports stationarity or non-stationarity.

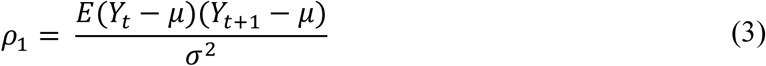

Equation (3) produces values ranging from -1 to 1. Non-stationarity time-series shows a slow decay in the ACF values while stationarity data exhibits rapid decay.

Next, regression modeling methods can be used to predict outcomes (e.g. mortality counts) in future years. Given a set of independent determinants (such as sex, age range and racial groups) an Ordinary Least Squares (OLS) regression can be used to predict outcomes, *y* using equation (4).

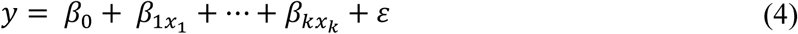

Where *β*_0_ is the intercept, *β*_1_ … *β*_*k*_, the coefficients representing the effect that the independent determinants have on *y*. The *ε* is an error term representing the difference between observed and predicted values. Notably, regression is sensitive to the relationship between independent variables. An independent variable should not be a linear combination of other variables (multicollinearity). This is especially common when categorical variables (common in health data) are encoded into numerical to fit in the model. Mathematically, a perfect multicollinearity occurs when the value of a third variable (*x*_3_) is exactly determined by the first two varibales (*x*_1_ and *x*_2_), see equation (5).

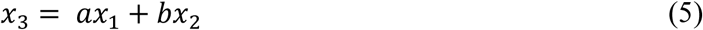

How well a regression model fits the data is defined using square of the residuals (*R*^2^). A large *R*^2^indicates that the independent variables explain the predictions better. A poor *R*^2^ implies that the regression model only explains a small portion of the variations. The independent variables might change based on location. The Global Moran’s *I* measure can be used to test whether the values are clustered, dispersed or randomly distributed (spatial autocorrelation). Therefore, Global Moran’s I (*I*) defined in equation (6) is useful in accurately modeling the data while considering spatial heterogeneity.

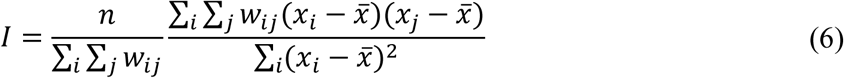

Where *x*_*i*_ is the value at location *i*, *w*_*ij*_ spatial weight between *i* and *j*. *n* is the number of features with a mean of *x̅*. If the value of *I* is greater than 0 (significant), spatial clustering exists while less than 0 indicates dispersion. An *I* value approximately equal to 0 implies spatial randomness in the variables. Considering a case of spatial clustering, Multiscale Geographically Weighted Regression (MGWR) analysis can be performed to describe the locations and how the relationship between predictors and the outcomes changes as defined in equation (7).

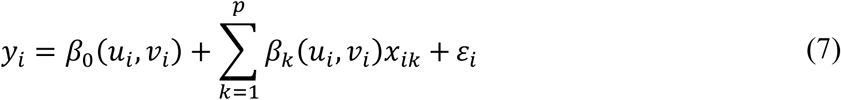

Where *u*_*i*_, *v*_*i*_ are the spatial coordinates of the place and *β*_*k*_(*u*_*i*_, *v*_*i*_) the location-specific coefficients. Therefore, each location has its regression represented by equation (8).

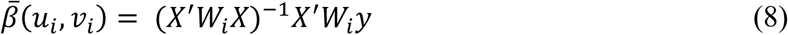

Generally, nearby points are related to each other thus hotspot analysis can be conducted to know which points influence each other using *z* score statistic expressed in equation (9).

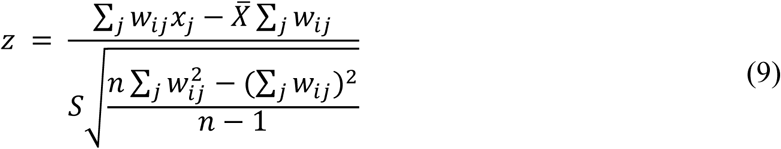

Where *x*_*j*_ is the value of the neighboring point *j*, *X̅* the global mean and *S* the standard deviation. A high positive z score indicates a hotspot while a high negative score represents a cold spot.

### 2.2. Application

Now that we have defined our theoretical methodology, we apply it to analyze 2005 to 2022 CDC PE mortality count data grouped by sex, age range and racial group. The data also specifies the state and the county Federal Information Processing Standard (FIPS) code where the death was registered. The data contained 72,704 datapoints. First, we group the mortality counts from 2005 to 2022 by sex, age range and race to detect increase or decrease in the cases by year. Next, we use the FIPs code to combine the CDC data with geographical layers of each of the US’s counties. Consequently, the result is a 2005 to 2022 mortality count for each of the US’s counties grouped by sex, age range and racial groups. We then conduct a chi-square test on the mortality counts by sex, age group and race to detect if the count distribution is independent across the counties.

Thirdly, we perform detailed time-series analysis of the mortality counts by calculating mean and variance within adjacent years to detect whether there is an increase or a decrease (stationarity test). We also fit an ACF to the dataset grouped by age range, racial group and sex.

Since, the data is grouped by sex, age range and race, we applied OLS to establish if future mortality counts can be predicted using these independent determinants. However, regression methods such as OLS are sensitive to multicollinearity. Moreover, knowing priori that the risk of PE mortality increases by age and considering the chi-test results, we use age as our main independent patient predictor. From past literature, studies have also shown that other environmental factors such as concentration of PM_2.5_ and poor air quality can cause blood coagulation and lead to deep vein thrombosis. Other factors such as poor walkability index and high poverty rates among neighborhoods can also influence behavior and the risk of PE. We therefore, use air quality (Environmental Protection Agency, 2022) and walkability index (Environmental Protection Agency, 2023) data from the US Environmental Protection Agency for each of the US counties. We also include the poverty and income data from the US Census Bureau (US Census Bureau, 2023). Thus, age range, PM_2.5_ concentration, walkability index and poverty rate are the main predictors for PE embolism in our OLS model.

Using the R^2^ and calculated residuals from the OLS model, we apply Global Morans I to test for spatial autocorrelation. We confirm a case of spatial clustering before applying MGWR to describe how the predictors affect the PE mortality outcomes across different counties. Lastly, we group the mortality counts into two periods (before 2015 and after 2015) to investigate emerging PE mortality hotspots across US counties.

## 3. Results

In this section, we report spatial-temporal results generated by following the methodology described in section 2. First, we report the temporal results to determine PE mortality count trends in the past 17 years. Next, we report the modeling regression results of PE mortality counts across US counties considering environmental and socioeconomic factors before presenting detailed hotspot analysis results.

### 3.1. Temporal results

According to Fig. 1A, PE mortality count for both males and females increased steadily over the study period. While both sexes had upward trajectory, the gap between their mortality count remains wide from 2005 to around 2015. The gap then narrowed post-2015 with a major jump in 2020, possibly due to the impact of Covid-19. Notably, the PE mortality count among females remained high compared to males across the study period.

**Fig. 1.**
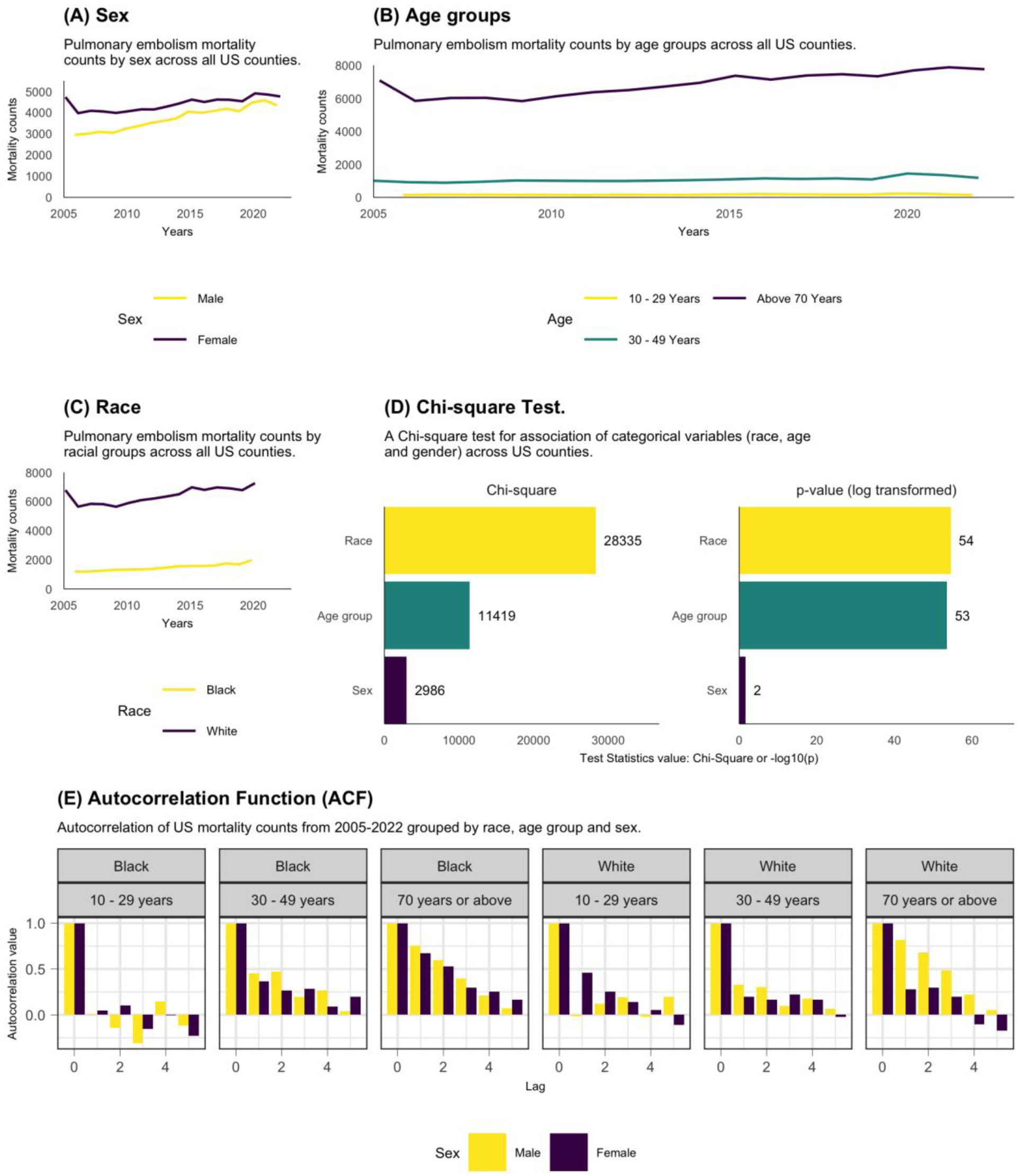
| Temporal analysis results. **A**, PE mortality counts by sex. **B,** PE mortality count by age ranges. **C,** PE mortality count results by racial groups. **D,** detailed stationarity test results for sex, age range and racial groups. **E,** ACF results grouped by all the demographic determinants.

**Fig. 2.**
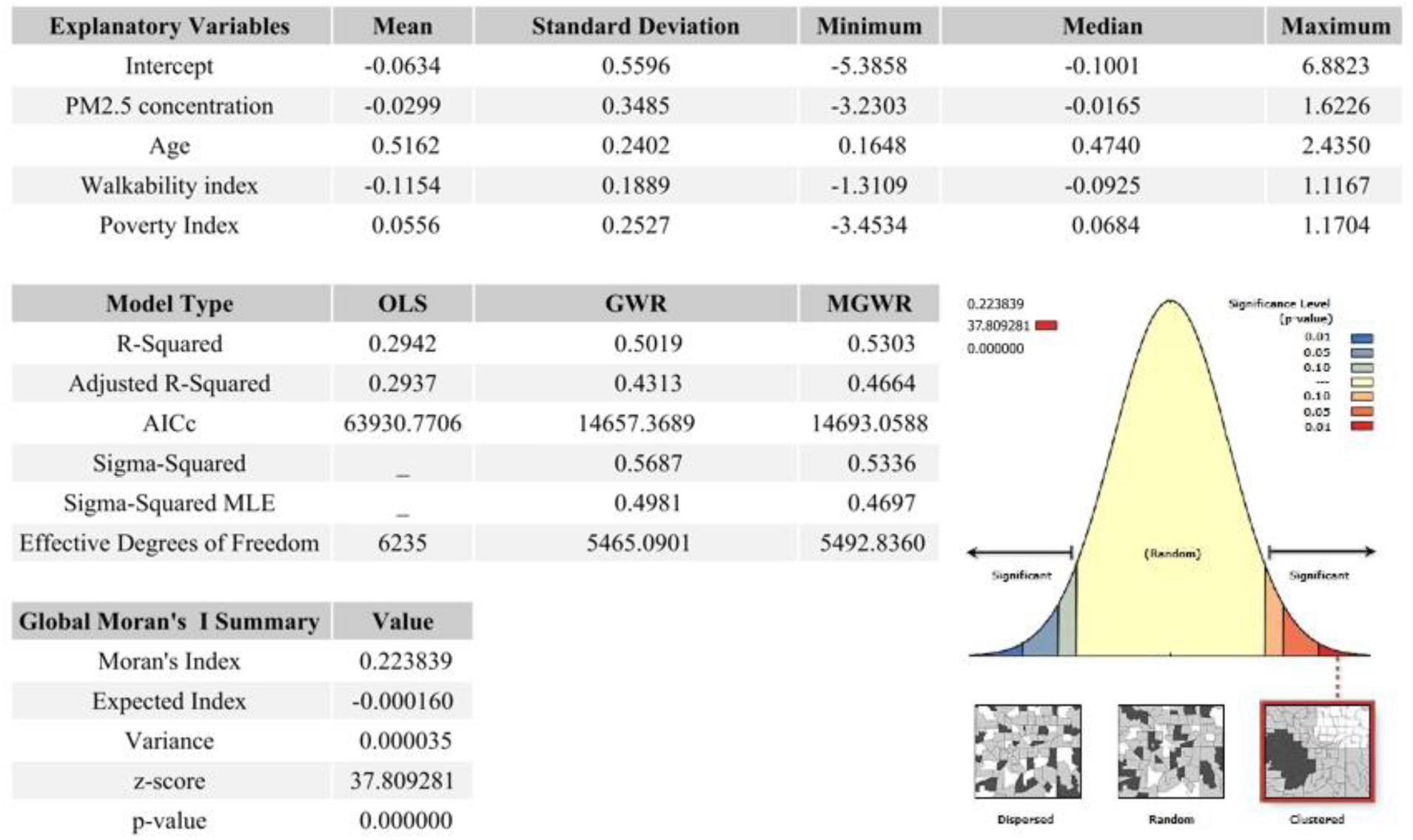
| Regression modeling results. Coefficient summaries, model diagnostics, and spatial autocorrelation statistics that indicate that PE mortality may be shaped by both socio-environmental factors under strong spatial non-stationarity.

In Fig. 1B, the trends in PE mortality counts are presented by age ranges. The PE mortality count is highest among individuals aged 70 years and above. The counts for 30-49 years group are relatively low and stable just like for 10-29 years group. The gap between age groups of 70 years and above is wide compared to the others (30-49 and 10-29 years) thus indicating advanced age as a dominant demographic risk factor for PE.

We also report mortality count by race across the study period (see Fig. 1C). The PE mortality count among White populations is consistently higher throughout the years compared to the Black populations. Whether these differences are a combination of population size effects or underlying disparities in PE risk, comorbidities, detection and access to care that warrant further adjusted analyses remain an open science question. Generally, the ACF results reported in Fig. 1E confirms an upward trend among races in all age-ranges and sex except for 10-29 years old among the Black population.

The chi-square test results reported in Fig. 1D assessing the association of PE mortality counts with sex, age range and racial groups shows that the latter has the strongest statistical association. Sex has the smallest but notable chi-square value. In general, p-values indicate that all associations are statistically significant.

### 3.2. Regression modeling results

To complement the descriptive and temporal results presented in section 3.1, we report regression modeling results. These results account for explanatory socio-environmental factors that may determine the risk of PE mortality across US counties tested under the assumptions of presence and absence of spatial-stationarity.

The descriptive statistics of demographic and possible socio-environmental factors contributing to PE mortality shows that age has the largest mean coefficient of 0.5162. Indeed, previous results had shown that PE mortality risk is associated with old age thus a significant predictor. Both PM_2.5_ concentration and walkability index have negative coefficients suggesting that more walkable environments may be associated with lower PE mortality. On the other hand, poverty rate index shows a modest mean, implying a potential socioeconomic gradient, although its effect is weaker compared to age. Notably, the wide ranges between minimum and maximum values across the variables suggest a case of spatial heterogeneity in relation to PE mortality.

Considering the explanatory variables, the spatially adaptive models performed better. For instance, the OLS model only explained 29% of the variance in PE mortality (R^2^ = 0.294). However, when allowing the covariates to operate at different spatial scales and using GWR and MGWR, the explanatory power increases to 50% (R^2^ = 0.502) and 53% (R^2^ = 0.53) respectively. The MGWR model, in particular, balanced complexity and interpretability by allowing multiscale spatial processes, which is appropriate given the heterogeneous demographic, environmental, and socioeconomic contexts across U.S. counties. Age for example showed the most positive association with PE mortality especially in Midwest counties of Mississippi, Kansas and Missouri. In these states the association between age and PE mortality was statistically significant in 96%, 91% and 90% of the counties respectively. Conversely, weaker or negative coefficients appeared in some western and northeastern counties, suggesting that the effect of age on PE mortality is moderated by regional factors.

The effect of environmental factors such as PM_2.5_ concentration also varies across counties especially among those with historically higher pollution burdens and population density such as Maryland, Virginia and North Carolina. The spatial clustering of positive PM_2.5_ effects supports the hypothesis that chronic air pollution contributes to thrombotic and cardiovascular risk, but in a geographically uneven manner. Although, the relationship between PM_2.5_ concentration is generally statistically weak in most counties, a higher positive coefficient was registered in 29%, 21% and 17% of counties in Maryland, Virginia and North Carolina respectively. In contrast, coefficients are weaker or negative across much of the western U.S. states, indicating either lower exposure levels or reduced sensitivity of PE mortality to PM_2.5_ in these areas.

The relationship between walkability index and PE mortality counts varies spatially. In coastal and densely populated areas, negative coefficients dominate implying that more walkable environments are associated with lower PE mortality, potentially through increased physical activity, reduced obesity, and improved cardiovascular health. However, positive coefficients in parts of the Midwest and South suggest that walkability may act as a proxy for urban density or other unmeasured risk factors in those regions. For instance, the relationship between walkability index and PE mortality counts was statistically significant in 39%, 33%, 24% and 15% of counties in Kansas, Maryland, Nebraska and Virginia respectively.

Lastly, the association of poverty index and PE mortality counts shows spatial variability especially in regions where it might exacerbate mortality through limited access to preventive care, delayed diagnosis, and higher prevalence of comorbid conditions. For instance, the relationship was more pronounced in Maryland and Nebraska States where it was statistically significant in 25% and 17% of the counties respectively.

These results indicate that PE mortality may not be governed by a single global process, but rather by spatially varying relationships that differ in strength and scale. The Global Moran’s I value of 0.224 provides further evidence of spatial clustering in PE mortality across the US counties.

### 3.3. Hotspot analysis results

In Fig. 4, we present the hotspot analysis results for the periods 2005-2015 and 2016-2022. In the period 2005-2015, the high mortality hotspot clusters (99% confidence) were concentrated in the South-Central (Texas, Arkansas, Oklahoma and Louisiana) and Southeastern states (Mississippi, Tennessee, Missouri). However, much of the cold spot clusters were concentrated in the western and northeastern states. These overall patterns persisted even in the period 2016-2022, though some states such as Kentucky, Alabama and Georgia had notable increase in PE mortality counts. For instance, 1.31%, 1.3% and 0.9% of counties in Kentucky, Alabama and Georgia respectively were PE mortality hotspots. In general, the PE mortality counts per 100,000 reduced between 2016-2022.

**Fig. 3.**
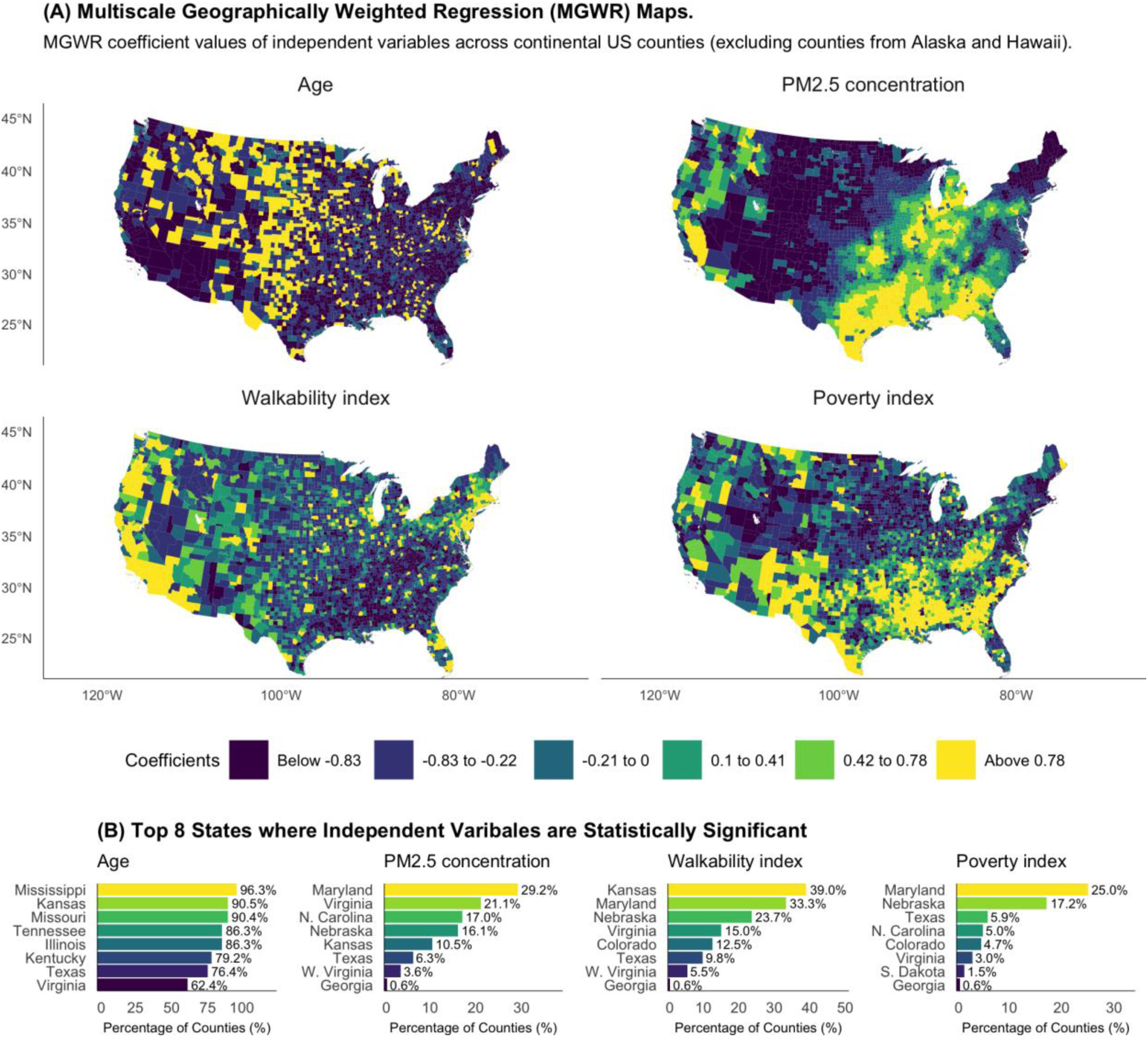
| Regression coefficients. **A**, Spatial distribution of MGWR coefficient estimates for age, PM2.5 concentration, walkability index, and poverty index. **B,** Top eight states with the highest proportions of counties in which each explanatory variable is statistically significant (p < 0.05) in the MGWR model.

**Fig. 4.**
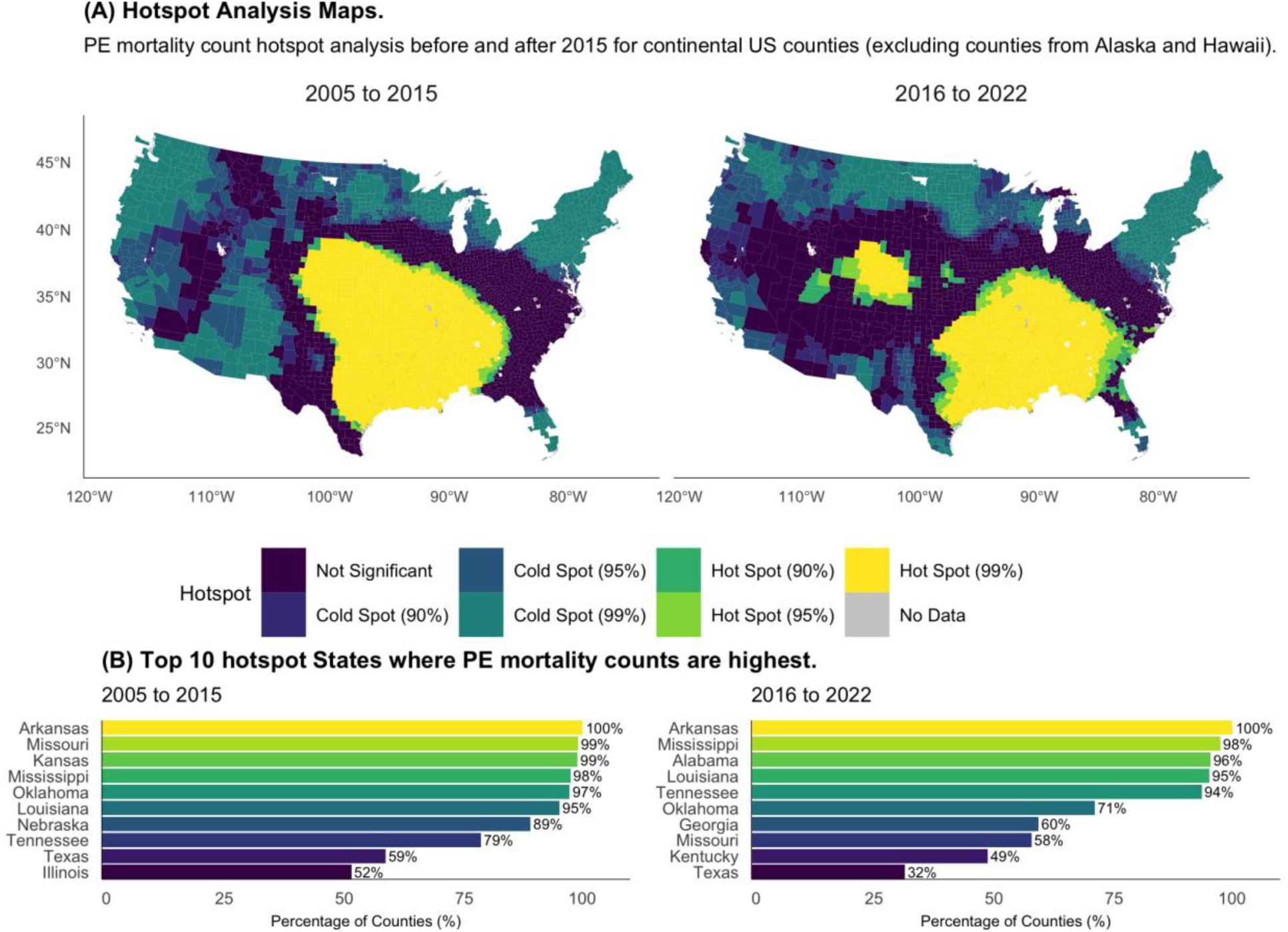
| Hotspot analysis results. **A**, Hotspot analysis of county-level PE mortality rates per 100,000 population for 2005–2015 and 2016–2022. **B,** Top ten states ranked by the percentage of counties classified as high-mortality hotspots in each study period.

We also present sex-stratified hotspot analysis results for the two period in Fig. 5. During 2005-2015, male PE mortality hotspots were concentrated in the states of Kansas (3.1%), Nebraska (2.8%), Mississippi (2.5%), Arkansas (2.5%), Oklahoma (1.6%), Missouri (1.4%), Louisiana (1.1%), Texas (0.6%), Tennessee (0.6%) and Colorado (0.5%). Most of the states remained male PE mortality hotspots in the period 2016-2022. Notably, the number of male PE mortality hotspots in Alabama increased significantly accounting for 1.2% of the counties which was not the case in 2005-2015.

**Fig. 5.**
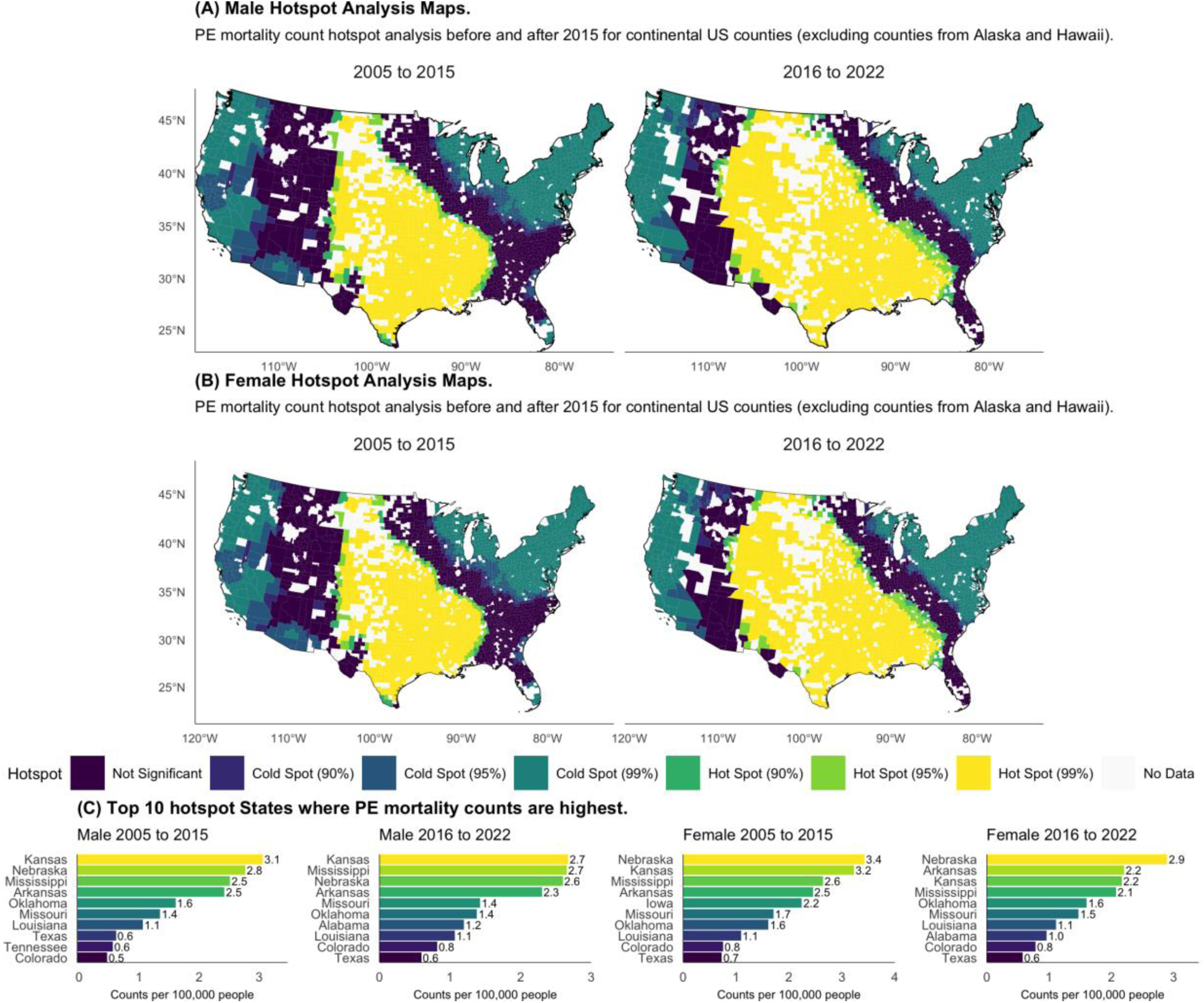
| Sex-stratified hotspot analysis. **A**, Male PE mortality hotspot maps for 2005–2015 and 2016–2022. **B,** Female PE mortality hotspot maps for 2005–2015 and 2016–2022. **C,** Top ten states ranked by the percentage of counties classified as high-mortality hotspots for males and females in each study period.

The female PE mortality hotspots broadly exhibited similar patterns but with notable differences in spatial extent and intensity. For instance, in 2005-2015, the hotpots were more spatially extensive including new states such as Iowa. Nebraska had the highest female PE mortality hotspots covering 3.4% of its counties while Texas had 0.7% among the hotspot states. During 2016-2022, the spatial extent reduced to 2.9% in Nebraska and 0.6% in Texas. However, Alabama emerged among the top ten states with the highest concentration of female PE mortality hotspots covering 1.0% of its counties.

## 4. Analysis and Discussion

Having presented trend, regression modeling and hotspot analysis results, we now undertake the discussion by revisiting the research questions previously articulated in section 1 as follows.

### 4.1. How do pulmonary embolism (PE) mortality counts vary over time across sex, age groups, and racial populations in the United States?

The results presented in this study shows that PE is not a transient phenomenon but rather a chronic and a fast-emerging challenge for health systems. The mortality counts across the demographic groups increase over time, consistent with growing recognition of PE as a major public health concern especially among the aging, and female population. Our analysis showed that age is a strong temporal determinant thus highlighting the importance of age-focused intervention strategies towards vulnerable older adults. Moving forward, policies that prioritize routine VTE risk assessment in elderly populations, improved post-hospitalization follow-up, and expanded access to anticoagulation management programs may yield substantial mortality reductions especially in predominant hotspot areas such as the Southern (Arkansas, Texas, Mississippi, Louisiana, Oklahoma) and Midwest (Nebraska, Missouri, Kentucky, Kansas, Tennessee) states.

The sex and racial differences in PE mortality patterns identified in this study further points to structural and systemic contributors. These may be in the form of health-seeking behavior, diagnostic delays, and access to timely care. Therefore, the strong statistical association between race and PE mortality suggests that equity-oriented interventions including standardized diagnostic pathways and improved access to preventive care in underserved communities are critical in addressing the PE challenge.

Overall, these temporal disparities indicate that effective PE control will require coordinated public health policies that integrate demographic risk stratification and targeted prevention efforts. Addressing PE as an epidemic demands both clinical and policy responses that extend beyond hospital settings to population-level risk reduction dictated by geography as identified in this study.

### 4.2. To what extent do demographic, socioeconomic, and environmental factors explain spatial variation in PE mortality across U.S. counties?

The regression modeling results strongly suggest that PE mortality rates across US counties are shaped by spatially heterogenous demographic, socioeconomic and environmental factors. Thus, global modeling approaches may severely understate these relationships. For instance, OLS model only explained 29% county-level variation in PE mortality compared to GWR and MGWR that explained nearly double the variation at 50% and 53% respectively.

As noted, age emerged as the most influential predictor of PE mortality, with the largest mean coefficient (0.5162) and statistically significant positive associations in a high proportion of counties, particularly in Mississippi, Kansas, Missouri, Tennessee, Illinois, Kentucky, Texas and Virginia. The results in these states provide empirical support for prioritizing regionally targeted geriatric PE prevention policies that may include mandatory VTE risk assessment protocols in hospitals, enhanced post-discharge anticoagulation monitoring, and expanded access to outpatient thromboprophylaxis programs in these high-risk regions. Thus, place-based risk stratification frameworks should guide resource allocation and clinical decision-making in counties with disproportionately aging populations as opposed to uniform state guidelines and policies.

Even though environmental and socioeconomic determinants displayed weaker global effects, we found meaningful spatial clustering that might have direct policy relevance. For instance, positive associations between PM_2.5_ concentration and PE mortality in 29% of counties in Maryland, 21% in Virginia, and 17% in North Carolina indicate that air-quality regulation and environmental health interventions may yield secondary benefits for PE outcomes in pollution-burdened regions. Also, the predominance of negative walkability coefficients in coastal and densely populated areas supports urban design and active-transport policies as potential population-level PE risk mitigation strategies. On the contrary, positive associations in parts of the Midwest and South caution against assuming uniform benefits of walkability without contextual evaluation. Lastly, the statistically significant poverty effects observed in up to 25% of counties in Maryland and 17% in Nebraska highlight the need for targeted investments in access to preventive care in socioeconomically vulnerable areas.

### 4.3. What are the spatial and temporal patterns of PE mortality hotspots across U.S. counties, and how does the distributions differ by period and sex?

Our analysis reveals that PE mortality is not randomly distributed across counties. Instead, the distribution of the mortality cases is geographically concentrated in clusters over time thus justifying the need to regionally frame PE public health problem. The PE mortality hotspots are concentrated in the counties of Arkansas, Mississippi, Kansas, Missouri, Oklahoma, Louisiana, Nebraska, Tennessee, and Texas repeatedly classified as statistically significant hotspots at the 99% confidence level. Although overall PE mortality rates per 100,000 declined during 2016–2022, in several states such as including Kentucky (1.3%), Alabama (1.3%), and Georgia (0.9%) indicating that aggregate national improvements may obscure worsening outcomes in specific regions. Thus, geographically targeted PE surveillance and intervention programs, rather than reliance on national or state averages to guide policy are necessary.

The sex-stratified hotspot patterns especially Male PE mortality in Kansas (3.1%), Nebraska (2.8%), Mississippi (2.5%), Arkansas (2.5%), and Oklahoma (1.6%) during 2005–2015 further highlight the importance of tailored policy responses. Moreover, the emergence of Alabama as a male hotspot state in 2016–2022, accounting for 1.2% of counties, signals shifting regional risk profiles that warrant proactive monitoring. On the other hand, female PE mortality hotspots were generally more spatially extensive in the earlier period, with Nebraska exhibiting the highest concentration (3.4% of counties), followed by Texas (0.7%). Although hotspot coverage among females contracted in 2016–2022 (2.9% in Nebraska and 0.6% in Texas), Alabama emerged again as a female hotspot state (1% of counties). Collectively, these results support sex-specific, place-based PE prevention strategies, to effectively reduce entrenched regional and demographic disparities in PE mortality.

## Conclusion

In this study we conducted a detailed spatial-temporal analysis of PE across counties between 2005 to 2022. Our study shows that PE mortality is not randomly distributed in space and time. The PE mortality hotspots are concentrated in the counties of Arkansas, Mississippi, Kansas, Missouri, Oklahoma, Louisiana, Nebraska, Tennessee, and Texas repeatedly classified as statistically significant hotspots at the 99% confidence level. When grouped by sex, our study reveals that Kansas (3.1%), Nebraska (2.8%), Mississippi (2.5%), Arkansas (2.5%), and Oklahoma (1.6%) are male PE mortality hotspots. On the other hand, majority of female PE deaths occurred in the counties of Nebraska (3.4%), Alabama (1%) and Texas (0.7%).

We also established that PE mortality rates are shaped by spatially heterogenous demographic, socioeconomic and environmental factors. Age emerged as the most influential predictor of PE mortality (mean coefficient of 0.52). It was the most statistically significant predictor with positive association in high proportion of counties of Mississippi, Kansas, Missouri, Tennessee, Illinois, Kentucky, Texas and Virginia. Our results thus provide empirical support for prioritizing regionally targeted geriatric PE prevention policies.

Importantly, the county-level analysis adopted in this study uncovers fine-scale geographic patterns that are usually obscured in state or national level analysis. By pinpointing counties with consistently elevated mortality risk, this work provides actionable evidence to support geographically tailored strategies aimed at reducing PE mortality. Overall, our findings indicate that reducing the threat of PE will not only require advances in clinical management but also place-based public health responses that addresses demographic and regional inequalities in risk exposure, healthcare access and outcomes. However, there is room for future research to integrate individual-level clinical data, healthcare infrastructure metrics, and environmental and socioeconomic determinants. Such a study will further clarify the mechanisms driving the observed spatial–temporal patterns and to inform more effective, equity-focused prevention and intervention strategies that is missing in this study.

## Data Availability

All data produced in the present study are available upon reasonable request to the authors

